# Development and validation of patient-level prediction models for adverse outcomes following total knee arthroplasty

**DOI:** 10.1101/2020.12.14.20240994

**Authors:** Ross D. Williams, Jenna M. Reps, Anthony G. Sena, Edward Burn, Ying He, Daniel R. Morales, David Culliford, Dahai Yu, Victoria Y. Strauss, Talita Duarte-Salles, Albert Prats-Uribe, Antonella Delmestri, James Weaver, William Sproviero, Danielle Robinson, Henry Morgan Stewart, Belay Birlie, Rafael Pinedo-Villanueva, Spyros Kolovos, Luis H. John, Ruth E. Costello, Michel van Speybroeck, Caroline O’Leary, Evan Minty, Thomas Falconer, Alison Callahan, Stephen Pfohl, Theresa Burkard, Jennifer Lane, Peter R. Rijnbeek, Patrick B. Ryan, Daniel Prieto-Alhambra

**Author notes:** These authors contributed equally to this work (Ross D. Williams, Jenna M. Reps). These authors contributed equally to this work (Patrick B. Ryan, Daniel Prieto-Alhambra).

## Abstract

**Background:** Elective total knee replacement (TKR) is a safe and cost-effective surgical procedure for treating severe knee osteoarthritis (OA). Although complications following surgery are rare, prediction tools could help identify those patients who are at particularly high risk who could then be targeted with preventative interventions. We aimed to develop a simple model to help inform treatment choices.

**Methods:** We trained and externally validated adverse event prediction models for patients with TKR using electronic health records (EHR) and claims data from the US (OPTUM, CCAE, MDCR, and MDCD) and general practice data in the UK (IQVIA Medical Research Database ([IMRD], incorporating data from The Health Improvement Network [THIN], a Cegedim database). The target population consisted of patients undergoing a primary TKR, aged ≥40 years and registered in any of the contributing data sources for ≥1 year before surgery. LASSO logistic regression models were developed for four adverse outcomes: post-operative (90-day) mortality, venous thromboembolism (VTE), readmission, and long-term (5-year) revision surgery. A second model was developed with a reduced feature set to increase interpretability and usability.

**Findings:** A total of 508,082 patients were included, with sample size per data source ranging from 1,853 to 158,549 patients. Overall, 90-day mortality, VTE, and readmission prevalence occurred in a range of 0.20%-0.32%, 1.7%-3.0% and 2.2%-4.8%, respectively. Five-year revision surgery was observed in 1.5%-3.1% of patients. The full model predicting 90-day mortality yielded AUROC of 0.78 when trained in OPTUM and yielded an AUROC of 0.70 when externally validated on THIN. We then developed a 12 variable model which achieved internal AUROC of 0.77 and external AUROC of 0.71 in THIN. The discriminative performances of the models predicting 90-day VTE, readmission, and 5-year revision were consistently poor across the datasets (AUROC<0.7).

**Interpretation:** We developed and externally validated a simple prediction model based on sex, age, and 10 comorbidities that can identify patients at high risk of short-term mortality following TKR. Our model had a greater discriminative ability than the Charlson Comorbidity Index in predicting 90-day mortality. The 12-feature mortality model is easily implemented and the performance suggests it could be used to inform evidence based shared decision-making prior to surgery and for appropriate precautions to be taken for those at high risk. The other outcomes examined had low performance.

**Funding:** This activity under the European Health Data & Evidence Network (EHDEN) has received funding from the Innovative Medicines Initiative 2 Joint Undertaking under grant agreement No 806968. This Joint Undertaking receives support from the European Union’s Horizon 2020 research and innovation programme and EFPIA. The sponsor of the study did not have any involvement in the writing of the manuscript or the decision to submit it for publication. The research was supported by the National Institute for Health Research (NIHR) Oxford Biomedical Research Centre (BRC). DPA is funded by a National Institute for Health Research Clinician Scientist award (CS-2013-13-012). TDS is funded by the Department of Health of the Generalitat de Catalunya under the Strategic Plan for Research and Innovation in Health (PERIS; SLT002/16/00308). The views expressed in this publication are those of the authors and not those of the NHS, the National Institute for Health Research or the Department of Health. The corresponding author had full access to all the data in the study and had final responsibility for the decision to submit for publication.

**Key Points:** *Question:* Is it possible to predict adverse events following total knee replacement?

*Findings:* Mortality was the only adverse event studied that we were able to predict with adequate performance. We produced a 12 variable prediction model for 90-day post-operative mortality that achieved an AUROC of 0.77 on internal test validation (Optum) and 0.71 when externally validated in THIN. The model also showed adequate calibration.

*Meaning:* Patients can now be presented with an accurate risk assessment for short term mortality such that they are well-informed before the decision for surgery is taken.

**Importance:** Total Knee Replacement is generally a safe, effective procedure that is performed on thousands of patients each year. However, a small number of those patients will experience adverse events. Due to the surgery’s elective nature, a well calibrated, high performing risk model could pre-emptively inform the patient and clinician decision making process and help to guide preventative treatment.

## Introduction

Total knee replacement is a common treatment for osteoarthritis (OA), with approximately 160,000 surgeries performed each year in the UK (1) and a predicted 1,065,000 surgeries in the US in 2020, rising to 1,429,000 in 2040 (2). This number is increasing, and it is predicted that by 2030 there will be 7.4 million people in the US living with a knee replacement. Total knee replacement (TKR) accounts for 90% of knee replacement surgeries (3), the rest are uni-compartmental knee replacements. TKR surgery is generally a safe procedure with fewer than 10% of patients experiencing post-operative complications. The adverse events can be separated into short term (90 days post operative) such as mortality and long term (5y post operative) such as revision surgery (5). Considering the example of short term mortality, if a patient is at high risk they may opt out of surgery as the long-term benefits may be less significant in comparison to the cost, both for the patients quality of life and the healthcare provider. Providing a risk prediction model to enable both clinicians and patients to identify risks of various post-operative complications will inform decision making regarding whether to opt for the surgery and to help target preventative interventions if surgery is performed.

Prediction models need to satisfy certain criteria to be clinically useful. Information must be readily available at the time of model implementation. For this study, that means all information used in the prediction model must be available pre-operatively. Further, models should be internally and externally validated demonstrating good performance. Current prediction model studies of post-operative outcomes after TKR have several limitations. In a systematic review of studies predicting post-operative infection after total joint replacement (6), most models were not externally validated and none were ready for clinical use likely due to issues of database interoperability. Moreover, some models were developed using data that were not routinely collected in observational data (e.g., floor of a patient’s bedroom, preoperative walking distance) and therefore validation of these models was infeasible using the data available in this study. Finally, most prediction models developed had not taken full advantage of all data available in medical records. For example, they only used a comorbidity index to predict mortality (7) instead of combining other patient characteristics, which would have likely improved the prediction of mortality (8). This knowledge gap demonstrates the need for novel patient-level prediction (PLP) models that can predict adverse events following TKR and should encapsulate all available pre-operative EHR data.

This study aimed to develop and validate models to predict the risk of post-operative complications following TKR, including venous thromboembolism (VTE), mortality, and readmission to hospital within 90 days, and 5-year revision using routinely collected observational healthcare records. Finally, we tested the TKR developed models in the UKR population to see if the performance transfers to this target population.

## Methods

### Study Design and Data Sources

This retrospective cohort study used observational healthcare databases from the UK and US. All datasets used in this paper were mapped into the Observational Medical Outcomes Partnership Common Data Model (OMOP-CDM) (10). The OMOP-CDM was developed for researchers to transform diverse datasets into a consistent structure and vocabulary. This enables analysis code and software to be shared among researchers which facilitates external validation of the prediction models.

#### Consent to publish

Each site obtained institutional review board approval for the study or used de-identified data and therefore the study was determined not to be human subjects research. Informed consent was not necessary at any site.

### Cohorts

#### Development Target Population Cohort

The target population for model development and validation contained patients undergoing TKR. The first recorded TKR procedure identified was considered the event of interest with the date of the surgery as the index date. Inclusion criteria required patients to have at least 1 year of continuous pre-index date recorded observation time. We excluded individuals below the age of 40, those with prior evidence of knee arthroplasty, knee fracture, knee surgery (except diagnostic procedures), rheumatoid arthritis, inflammatory arthropathies, or septic arthritis at any time before the index date. Patients with spine, hip, or foot pathology observed in the 365 days before index date were also excluded.

#### Validation Target Population Cohort

Uni-compartmental knee replacement (UKR) is an alternative to TKR surgery and we were interested to determine whether the models developed on the TKR patients were transportable to UKR patients. The UKR cohort was defined as patients with a UKR procedure with the initial procedure date corresponding to the index date. Inclusion and exclusion criteria were the same as in the TKR cohort. This cohort was used to assess performance transfer.

The target cohorts for TKR and UKR are available at:

TKR: http://www.ohdsi.org/web/atlas/#/cohortdefinition/1769719

UKR: http://www.ohdsi.org/web/atlas/#/cohortdefinition/1769730

#### Outcome Cohorts

The post-operative complications assessed were an occurrence within 90-days of VTE, all-cause readmission, all-cause mortality and within 5-year revision. Not all databases contained sufficient information to use every outcome. For example, THIN is a general practice database and does not contain full information on hospitalization thus we were unable to perform the readmission analysis.

1. VTE was identified based on observing a diagnosis code of either deep vein thrombosis or pulmonary embolism. Available at:http://www.ohdsi.org/web/atlas/#/cohortdefinition/1769735
2. Readmission was defined as the first inpatient or emergency room visit for any cause after knee replacement and was assessed in the OPTUM claims database. Available at: http://www.ohdsi.org/web/atlas/#/cohortdefinition/1769734
3. Mortality was identified based on records of date of death. Available at:http://www.ohdsi.org/web/atlas/#/cohortdefinition/1769731
4. Implant survival was assessed in terms of knee arthroplasty revision, with instances of revision identified based on observing a procedure code for revision. Available at:http://www.ohdsi.org/web/atlas/#/cohortdefinition/1769726

#### Time at risk

We investigated two different time-at-risk (TAR) periods to predict short and long-term outcomes. Patients were considered at risk for VTE, readmission, and mortality from the day after surgery up until day 90. For revision, we predicted the outcomes occurring the day after surgery until 5 years.

#### Candidate Predictors

We derived 89,031 candidate predictors from the observational healthcare data that existed on or prior to the target index date (TKR surgery date). These variables were demographics, binary indicators of medical events and counts of record types. The demographics were gender, binary indicators of age in 5 year groups (40-45, 45-50,…,95+) and month of the target index date. We created binary indicator variables for medical events based on the presence or absence of each concept for a patient corresponding to the OMOP-CDM clinical domains of conditions, drugs, procedures or measurements. For conditions we created binary predictors using the 30 days and 365 days prior to index date. For example, there exists one covariate for each of ‘Diabetes mellitus’, ‘Hypertensive disorder’, and ‘Hypercholesterolemia’ (and all other diseases), based on the occurrence of a diagnosis code for each condition in the 365 days or 30 days preceding the index date. For drugs we created binary indicator variables for drug records that occurred within 30, 365, 1095 days and all time prior to target index date. We also created count covariates representing how many visits a patient had in the 365 days and 1095 days prior to the target index date. For example, if a patient had 3 inpatient visits in the 365 days prior to index then their value for the variable “inpatient visit count −365 to 0 day prior to index” would be 3. Existing risk scores (CHADS2, CHA2DS2VASc (both stroke risk models), Diabetes Complications severity index, Charlson Comorbidity Index) using all data prior to index were also calculated and used as candidate predictors.

#### Methodology for model development and validation

The study was initially conducted using the THIN, OPTUM, MDCD, MDCR and CCAE datasets. For each suitable dataset we developed models predicting the 5 adverse events in the TKR target population. Mortality is better captured in OPTUM and THIN, so we developed two 90-day mortality models. Readmission was only captured in the claims data (OPTUM, MDCD, MDCR and CCAE), so we developed four 90-day readmission models. VTE and revision are captured in all five datasets, so five models were developed for each of these outcomes. This resulted in a total of 16 models. We then utilized the interoperability of the OMOP-CDM to perform a network study that enabled us to externally validate models that showed adequate internal discriminative ability.

Model development followed the framework for the creation and validation of PLP models presented in Reps et al (12), we used a person ‘train-test split’ method to perform internal validation. In each development cohort, the random split sample (‘training sample’) containing 75% of patients was used to develop the prediction models and the remaining 25% of patients (‘test sample’) was used to validate the risk scores. We trained models using LASSO regularised logistic regression, using a 3-fold cross validation technique in the training sample to learn the optimal regularisation hyper parameter through an adaptive search (13).

We assessed the performance of the model in terms of discrimination and calibration. Discrimination assesses how well the model can distinguish between patients who do or do not go on to experience the outcome and calibration which assesses whether the predicted risks are in alignment with the observed risks. Discrimination was measured using the Area Under Receiver Operator Characteristic Curve (AUROC). The AUROC indicates the probability that a randomly selected patient with an outcome during the TAR will be assigned a higher risk by the model than a randomly selected patient without the outcome during the TAR. We generally consider an AUROC of greater than 0.70 to be a reasonable candidate for external validation. The model calibration was presented in a plot to examine agreement between predicted and observed risks across deciles of predicted risk. Calibration assessment is then performed visually rather than using a statistic or numeric value as this provides an impression of the direction and scale of miscalibration (14). The sensitivity (how many people who will experience the outcome will be detected by the model) and positive predictive value (the probability that a patient detected by the model will experience the outcome) at various thresholds are also presented. Summary statistics were reported from the test samples.

We performed external validation (15) in which we applied the final prediction models to each suitable dataset not used to develop the model (suitability defined as containing the required information). For example, some of the datasets studied did not record death well and as such were not suitable for a mortality model. We examined the external validation using AUROC and calibration in the same way as internally.

#### Making the models more parsimonious

When using a data driven approach to model development, generally the final models contain a large number of covariates. The expectation was that for the models created in this study, this would be the case. The full model assesses what is in principle the best possible performing model, which can be demonstrated to be both transportable and robust when externally validated. However, the large number of covariates creates an implementation barrier and to some extent a barrier to the understanding of the model.

We therefore created models that could be candidates for the clinical implementation by performing further analyses in order to reduce the number of features in the final model (improving parsimony). This analysis investigated what the performance loss is when using fewer covariates.

The approach involves analyzing the covariates selected by the final model and then using clinical expertise to attempt to combine multiple of these covariates, that correspond to a similar illness, into a single covariate. Often the LASSO logistic regression model includes multiple covariates which are potentially clinically related, for example a model might select the same condition occurrence but in different time periods predating the index date (e.g., ‘diabetes −30 days to 0 days prior to index’ and ‘diabetes −365 days to 0 days prior to index’). These could be simplified to a covariate saying only “Patient had diabetes any time prior to index”, rather than multiple covariates specifying the specific time frame of the occurrence.

Finally, once the new aggregate covariates have been identified, the analysis was run again on the original databases using only the aggregate covariates to create a parsimonious model with a reduced number of covariates that aims to perform similarly to the full model. The procedures for developing both the full and parsimonious models will be identical except for the covariates. Definitions of the aggregated covariates are available in Supplement 1.

We used R (version 3.5.1) and the Patient-Level Prediction package for all statistical analyses. This study was conducted and reported according to the Transparent Reporting of a multivariate prediction model for Individual Prediction or Diagnosis (TRIPOD) guidelines (16). We were able to perform external validation of the models by leveraging the OHDSI community network and standardization. The OHDSI network is a community of researchers who have converted their medical record data into the OMOP-CDM which allows for rapid model dissemination and validation together with the creation of the Patient-Level Prediction R package. All the analysis code used for the development for the models are available on github at https://github.com/OHDSI/StudyProtocolSandbox/tree/master/mortalityTRK as well as the developed mortality models themselves for external validation at: https://github.com/ohdsi-studies/TkrPredictSimple

## Results

The target population ranged from 1,853 (MDCD) to 158,549 (CCAE) across the five datasets used to develop the prediction models. In general, the incidences of the various adverse events following TKR were similar across the datasets with 90-day mortality occurring in 0.20-0.23% of patients, 90-day readmission occurring in 7.07-8.98% of patients, 90-day VTE occurring in 2.20-4.74% of patients and 5-year revision occurring in 1.48-3.13% of patients (Table 1).

**Table 1.**
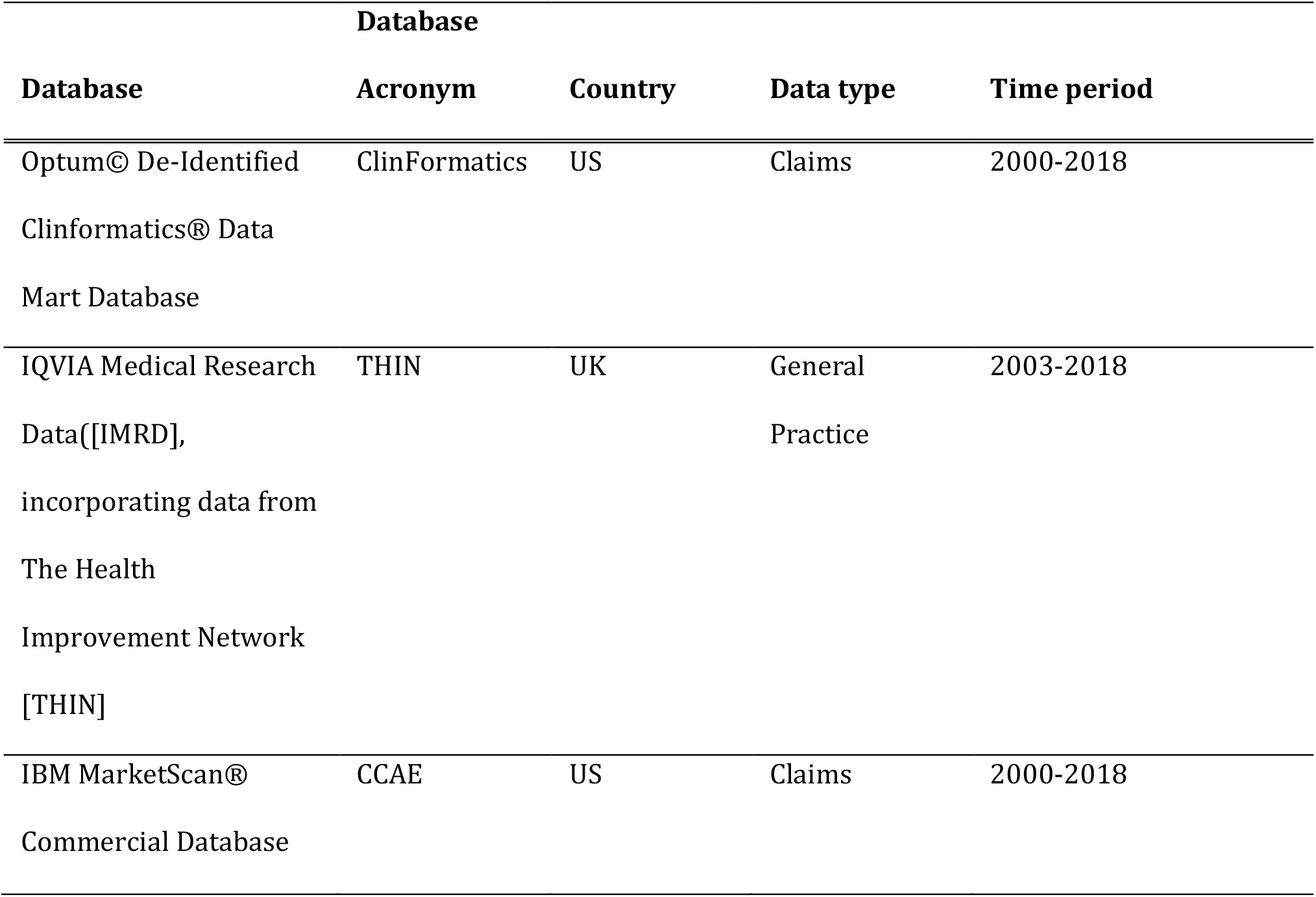
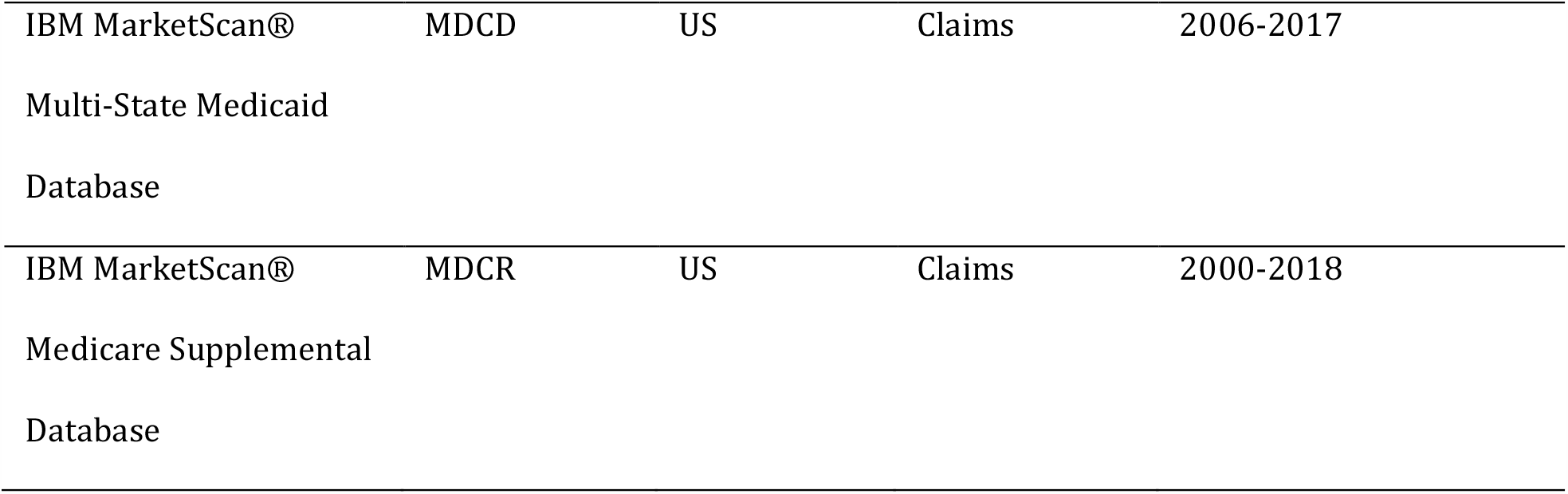
Data sources formatted to the Observational Medical Outcomes Partnership Common Data Model (OMOP-CDM) used in this research (data type: claims, electronic health/medical records (EHR/EMR), general practitioner (GP))

Out of the 16 models developed in this study, only the 90-day mortality model trained using OPTUM or THIN and the 90-day readmission model trained using MDCD obtained internal AUROC around 0.7 or higher (Table 1). When investigating the generalizability of the models, the external validation of the 90-day readmission was poor (AUROC between 0.58-0.60). The external validation of the 90-day mortality models developed on OPTUM and THIN ranged between 0.68 to 0.86 and are presented in Table 2.

**Table 2.** TKR target and outcome population sizes and the internal AUROC achieved

| Dataset | Target Population | 90-day mortality |  | 90-day readmission |  | 90-day VTE |  | 5-year revision |  |
| --- | --- | --- | --- | --- | --- | --- | --- | --- | --- |
|  |  | Size | AUROC | Size | AUROC | Size | AUROC | Size | AUROC |
| OPTUM | 152,665 | 353<br>(0.23%) | 0.78 | 10,792<br>(7.07%) | 0.63 | 6,723<br>(4.40%) | 0.62 | 3,134<br>(2.05%) | 0.63 |
| THIN | 40,950 | 81<br>(0.20%) | 0.68 | NA | NA | 900<br>(2.20%) | 0.61 | 607<br>(1.48%) | 0.61 |
| CCAE | 158,549 | NA | NA | 12,145<br>(7.66%) | 0.69 | 5,848<br>(3.69%) | 0.61 | 3,844<br>(2.42%) | 0.60 |
| MDCR | 136,467 | NA | NA | 12,259<br>(8.98%) | 0.66 | 6,488<br>(4.75%) | 0.56 | 2,716<br>(1.99%) | 0.63 |
| MDCD | 15,292 | NA | NA | 1,230<br>(8.04%) | 0.60 | 559<br>(3.66%) | 0.61 | 478<br>(3.13%) | 0.63 |

The OPTUM 90-day mortality model performed better than the THIN 90-day mortality model both internally and across the external validation (Table 2). The OPTUM 90-day mortality model achieved a slightly increased performance (AUROC 0.69) in the THIN dataset compared to the internal validation of the THIN developed model (AUROC 0.68).

The models and performance on the test and external validation sets are available to explore interactively at http://data.ohdsi.org/TKROutcomesExplorer/

We further validated the full Optum trained 90-day mortality TKR model in the UKR cohorts in the OPTUM dataset. This achieved an AUROC of 0.94 which suggests that performance is transferable between the two surgeries. However due to the low number of UKR surgeries that are carried out, there were only 11 deaths so the performance is possibly unreliable.

The prevalence of a selection of covariates included in the 90-day mortality model developed using OPTUM, when assessed in multiple databases can be found in Appendix 1.

This analysis shows that the covariate prevalence varies between the different databases, suggesting the databases have different underlying characteristics. As the models maintain performance despite these differences, it suggests that the model is robust to variability in the distribution of the covariates.

When the number of patients with a covariate is fewer than 5 for privacy reasons this is marked as 0.

Increasing the parsimony of the 90-day Optum mortality model was then performed. The aggregate covariates are detailed in Appendix 1. This model is detailed in Table 3.

**Table 3.**
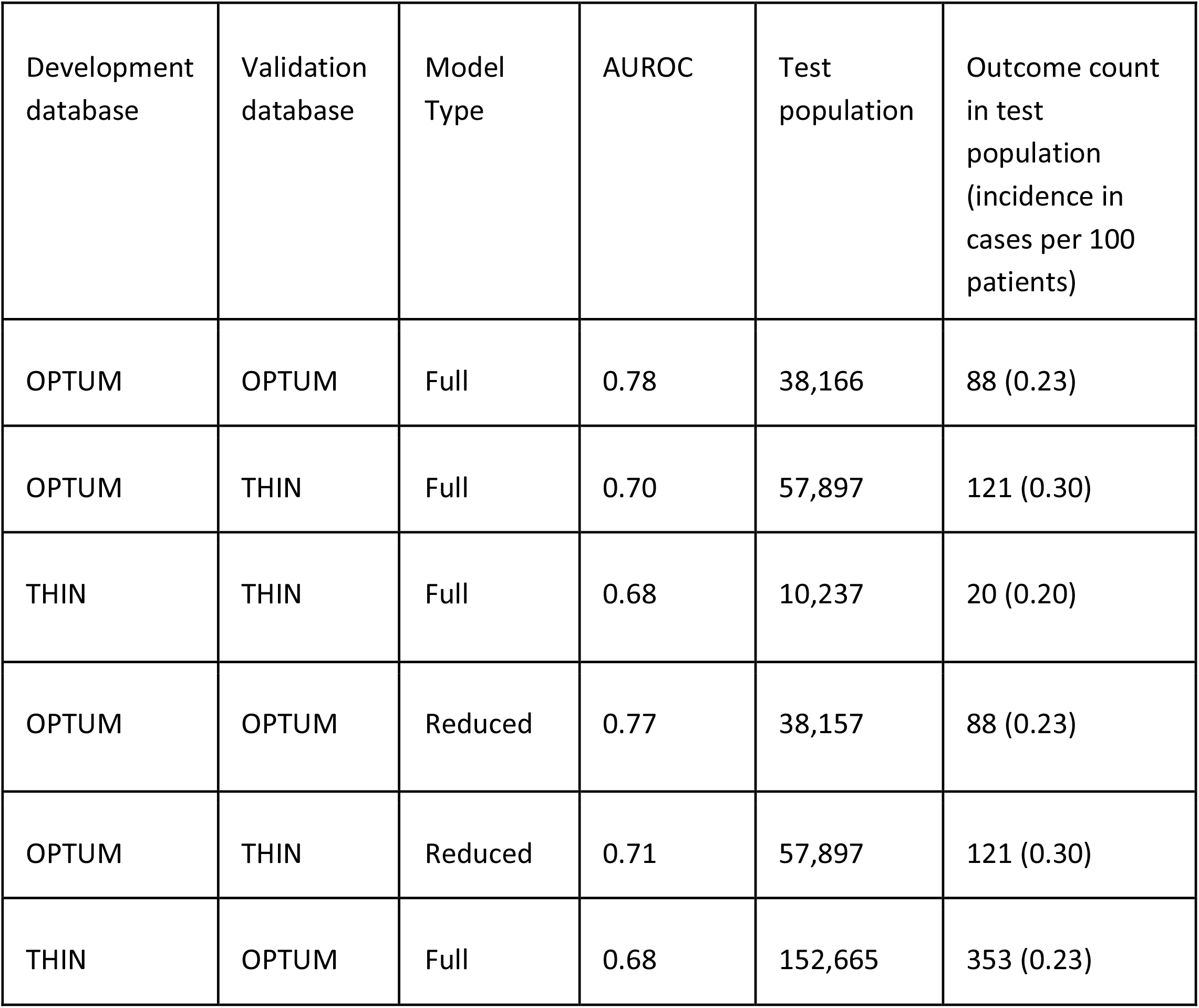
The external validation of the 90-day mortality models

**Table 4.**
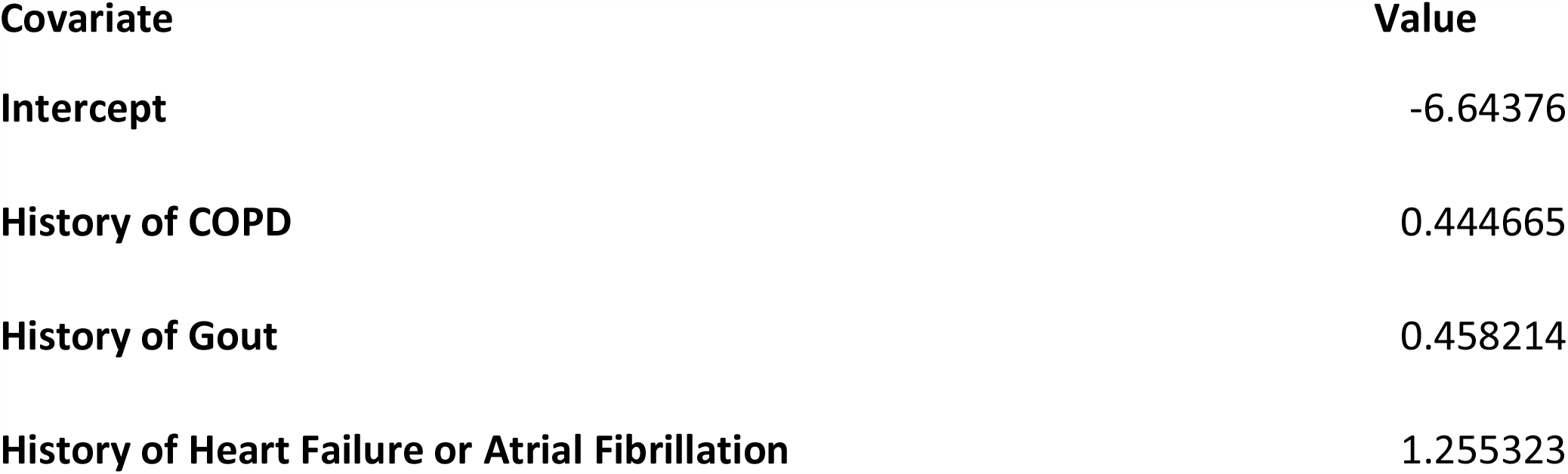
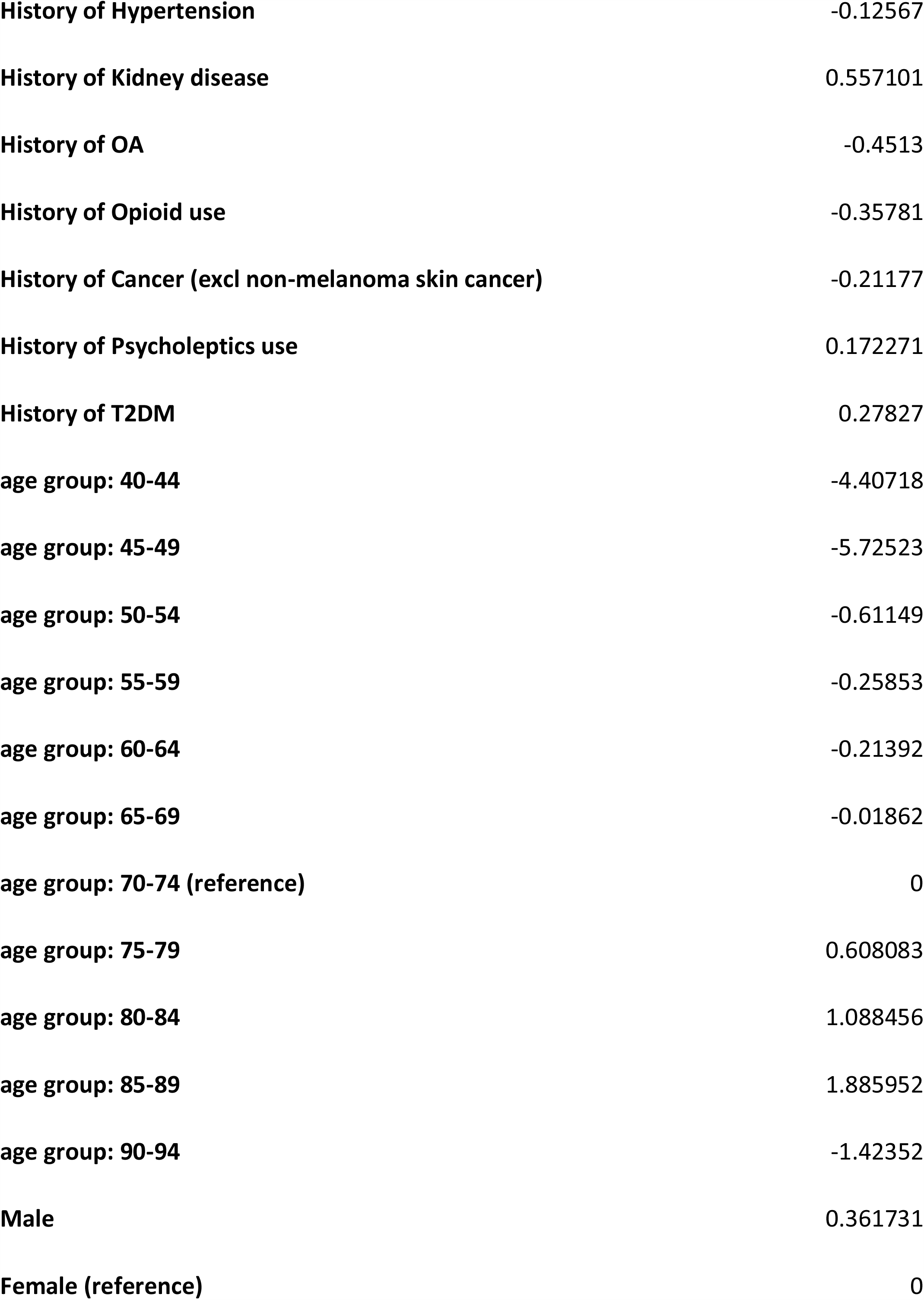
The covariates and values for the parsimonious 90-day mortality model

**Table 5.** Optum simple model performance at various thresholds

| Finding people with the outcome.... |  |  |  |  |
| --- | --- | --- | --- | --- |
|  | Prediction threshold | Sensitivity of death | Specificity of death | Positive predictive value |
| If you choose by prediction **threshold**... |  |  |  |  |
|  | 0.001 | 0.86 | 0.50 | 0.004 |
|  | 0.005 | 0.40 | 0.91 | 0.009 |
|  | 0.01 | 0.23 | 0.97 | 0.017 |
| If you choose by sensitivity... |  |  |  |  |
|  | 0.0038 | 0.50 | 0.86 | 0.008 |
|  | 0.0014 | 0.75 | 0.63 | 0.005 |
|  | 0.0008 | 0.90 | 0.36 | 0.003 |
| If you choose by specificity.... |  |  |  |  |
|  | 0.0048 | 0.43 | 0.90 | 0.009 |
|  | 0.0087 | 0.30 | 0.95 | 0.014 |
|  | 0.020 | 0.13 | 0.99 | 0.029 |

When the analysis was performed with these new covariates, the AUROC achieved was 0.77 internally in Optum and 0.71 in THIN. The calibration plot for the internal validation and the THIN validation are presented in Figure 1.

**Figure 1.**
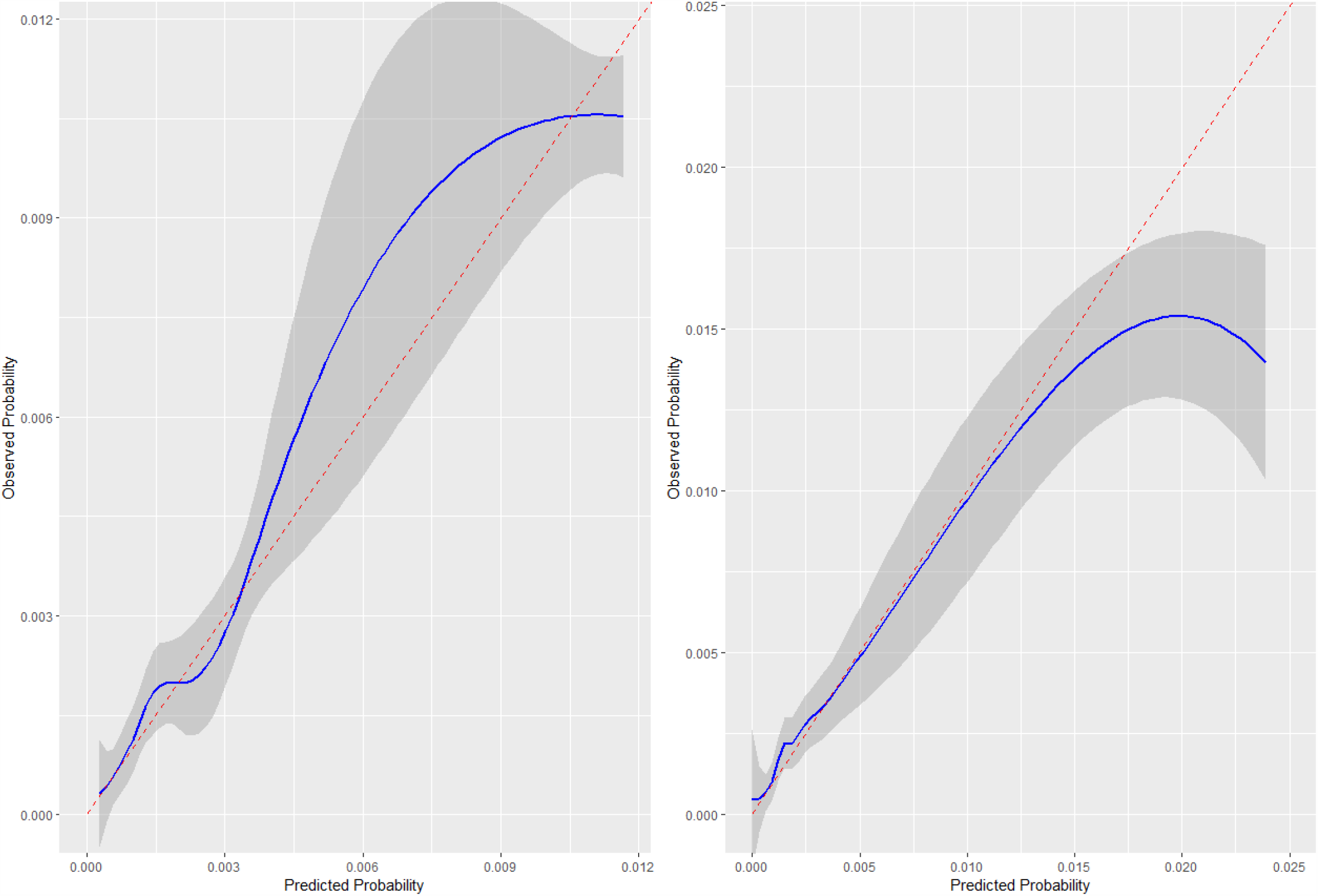
Calibration plot showing the calibration in Optum (left) and THIN(right)

## Discussion

This study created and evaluated prediction models for short-term (90-day) mortality, VTE, readmission, and long-term (5-year) risk of implant revision, of TKR in retrospective cohorts derived from UK and US databases.

The main finding of this study is the predictability of post-operative 90-day mortality following TKR. The AUROC of LASSO logistic regression model was found to be 0.78 (OPTUM). Validating this model against the other databases resulted in AUROC values of 0.69 (THIN) indicating that the model is fairly robust. For the 90-day mortality OPTUM model, 102 of 89,031 candidate variables were selected into the final model. The full model is available in Appendix 2. Some of the predictors for mortality in TKR patients included being aged over 75, existing cardiovascular disease, kidney disease, recent hospitalization, chronic obstructive pulmonary disorder, and being male. This high number of features presents a barrier to implementation in clinics and, as such, the parsimonious model, which achieved AUC of 0.77 (internal) and 0.71 (THIN), is preferred. The calibration of this model is presented in Figure 1 and shows that, for the majority of patients, the model is well calibrated internally with the ideal line always appearing within the confidence interval. For the external validation in THIN, in the low to medium risk range the model is well calibrated however for patients at higher risk with some underestimation of risk in the highest risk groups. For example, a predicted risk of 0.02 corresponds to an observed risk of 0.015. Both plots were made over the prediction region that contained patients and as such there is no information available on the calibration of patients at higher predicted risk. This is likely to be a small number in practice.

The desired operating characteristics when applying the parsimonious OPTUM 90-day mortality model to classify patients into those who will die and those who will not within 90 days of the surgery can be picked based on the prediction threshold, see Table 3. For example, if a reader wished to identify patients at risk of mortality after the surgery with a specificity of 0.95 (i.e. 5% of those who don’t die were predicted as dying), then they can scroll down the “Specificity of death” column until they reach the row with the 0.95 value and then they can see this would correspond to identifying patients assigned a risk of 0.0087 or higher by our model and the sensitivity would be 0.30 (i.e. 30% of patients are correctly identified as dying). Alternatively, if the reader wishes to identify 50% of those who will die shortly after surgery, they would scroll down the “Sensitivity of death” column until they reach the row with the value 0.50 which would show they need to identify patients with a predicted risk of 0.0038 or greater. This would achieve a specificity of 0.86. One way of interpreting this threshold is to consider patients who have extremely high or low risks. If the threshold is set at 2% then patients who have a risk higher than this (a risk an order of magnitude greater than the average) will be predicted to die following surgery. In our example this would identify approximately 10% of the patients who die following surgery whilst flagging around 1% of the population. At the other end patients who have an extremely low predicted risk can also be identified as such with good confidence.

In contrast to previous studies, the focus of this research was to develop the best performing predictive model on basis of all clinical and demographic data recorded in the observational databases and to then assess how close to this performance a reduced feature set model could come.

Hunt et al. report an incidence of mortality (0.37%) in their study on 45-day mortality following knee replacement surgery(17). This is high compared with our reported incidence of mortality, which could be due to the limitation of the mortality capture in the databases studied. The low incidence of death (around 0.2%) following TKR necessitates large datasets with accurate recording of mortality. The reported 90-day mortality predictive model may be used as a complementary element for screening of high-risk patients and better preparation before surgery. It could also allow the patient and clinician to be better informed about the potential benefit-risk of elective TKR. Given that we considered all-cause mortality the mortality is not necessarily caused by the TKR, however if the patient is deemed to be at a high risk of mortality in the 90-day postoperative period then the surgery is still likely inadvisable due to the costs to both the patient and the healthcare system without providing benefit.

Concerning the non-mortality outcomes investigated in this paper, we were unable to produce high performing risk models using a data driven approach. This suggests that it is difficult to identify patients at high risk in the data used in this study. As such it is unwise to base any advice for patients on a perceived high or low personal risk of having the outcome using the models developed. Until accurate models for these outcomes are developed.

Limitations of this study include the low number of outcomes in some of the analyses meaning that estimates are potentially unreliable, as well as potential misclassification of covariates in the data. The recording of death in the THIN is very reliable but in Optum is known to be specific but lacking some sensitivity due to a change in the reporting in 2013 when death reporting of death stopped being mandatory. This could lead to an underestimation of the number of deaths following a TKR in this study.

Limitations of the phenotypes include: i) assuming 5-year revision is for identified TKR ii) there is a potential contamination issue in the TKR cohort as prior to ICD-10 coding, TKR cohorts will have UKR cases as the same ICD procedure code was valid for both iii) if a patient were to have bilateral TKR we would only include the first surgery in our target cohort and the second would be excluded.

A major strength of this study is the external validation of prediction models. Typically, it takes 3 or more years for a prediction model to be externally validated due to reasons such as different coding systems in different databases causing difficulties in implementation (12). In this study, we presented multiple external validations, which demonstrated its robustness and transportability. Furthermore, external validation clearly demonstrated the quality of the developed model. The low number of features of this model is a significant advantage to implementation.

In conclusion, we developed and performed external validation for a 90-day mortality after a TKR prediction model that has both good discrimination performance and calibration which are maintained across the external validation. Thus, we believe that it is a strong candidate for impacting clinical decision making.

## Data Availability

Due to patient privacy concerns no patient level data is available however all aggregated results have been made available

## Acknowledgements

The authors would like to thank the OHDSI community for their contributions to the tools used for this analysis. The authors would also like to thank Paloma O’Dogherty Cordero who helped to organise the Oxford OHDSI study-athon, from which this study emerged. IQVIA Medical Research Data incorporates data from The Health Improvement Network, THIN. THIN is a registered trademark of Cegedim SA in the UK and other countries. Reference made to the THIN database is intended to be descriptive of the data asset licensed by IQVIA. This work uses de-identified data provided by patients as a part of their routine primary care.

## Author contributions

All authors made substantial contributions to the conception or design of the work; RW, PR, DPA and JL constructed the aggregate covariates DPA and PBR led the acquisition of the data; all authors were involved in the analysis and interpretation of data for the work; All authors have contributed to the drafting and revising critically the manuscript for important intellectual content; all authors have given final approval and agree to be accountable for all aspects of the work.

## References

1. Registry NJ. National Joint Registry: National Joint Registry for England, Wales and Northern Ireland; 15th Annual Report. 2018.

2. Singh JA, Yu S, Chen L, Cleveland JD. Rates of Total Joint Replacement in the United States: Future Projections to 2020-2040 Using the National Inpatient Sample. J Rheumatol. 2019;46(9):1134–40.

3. Maradit Kremers H, Larson DR, Crowson CS, Kremers WK, Washington RE, Steiner CA, et al. Prevalence of Total Hip and Knee Replacement in the United States. J Bone Joint Surg Am. 2015;97(17):1386–97.

4. Pearse RM, Moreno RP, Bauer P, Pelosi P, Metnitz P, Spies C, et al. Mortality after surgery in Europe: a 7 day cohort study. Lancet. 2012;380(9847):1059–65.

5. Springer BD, Cahue S, Etkin CD, Lewallen DG, McGrory BJ. Infection burden in total hip and knee arthroplasties: an international registry-based perspective. Arthroplast Today. 2017;3(2):137–40.

6. Kunutsor SK, Whitehouse MR, Blom AW, Beswick AD. Systematic review of risk prediction scores for surgical site infection or periprosthetic joint infection following joint arthroplasty. Epidemiol Infect. 2017;145(9):1738–49.

7. Inacio MCS, Pratt NL, Roughead EE, Graves SE. Evaluation of three co-morbidity measures to predict mortality in patients undergoing total joint arthroplasty. Osteoarthritis Cartilage. 2016;24(10):1718–26.

8. Konopka JF, Hansen VJ, Rubash HE, Freiberg AA. Risk assessment tools used to predict outcomes of total hip and total knee arthroplasty. Orthop Clin North Am. 2015;46(3):351-62, ix-x.

9. Beard D, Price A, Cook J, Fitzpatrick R, Carr A, Campbell M, et al. Total or Partial Knee Arthroplasty Trial - TOPKAT: study protocol for a randomised controlled trial. Trials. 2013;14:292.

10. Overhage JM, Ryan PB, Reich CG, Hartzema AG, Stang PE. Validation of a common data model for active safety surveillance research. J Am Med Inform Assoc. 2012;19(1):54–60.

11. Blak BT, Thompson M, Dattani H, Bourke A. Generalisability of The Health Improvement Network (THIN) database: demographics, chronic disease prevalence and mortality rates. Inform Prim Care. 2011;19(4):251–5.

12. Reps JM, Schuemie MJ, Suchard MA, Ryan PB, Rijnbeek PR. Design and implementation of a standardized framework to generate and evaluate patient-level prediction models using observational healthcare data. J Am Med Inform Assoc. 2018;25(8):969–75.

13. Suchard MA, Simpson SE, Zorych I, Ryan P, Madigan D. Massive parallelization of serial inference algorithms for a complex generalized linear model. ACM Trans Model Comput Simul. 2013;23(1).

14. Iqbal J, Vergouwe Y, Bourantas CV, van Klaveren D, Zhang YJ, Campos CM, et al. Predicting 3-year mortality after percutaneous coronary intervention: updated logistic clinical SYNTAX score based on patient-level data from 7 contemporary stent trials. JACC Cardiovasc Interv. 2014;7(5):464–70.

15. Reps JM, Williams RD, You SC, Falconer T, Minty E, Callahan A, et al. Feasibility and evaluation of a large-scale external validation approach for patient-level prediction in an international data network: validation of models predicting stroke in female patients newly diagnosed with atrial fibrillation. BMC Med Res Methodol. 2020;20(1):102.

16. Moons KG, Altman DG, Reitsma JB, Ioannidis JP, Macaskill P, Steyerberg EW, et al. Transparent Reporting of a multivariable prediction model for Individual Prognosis or Diagnosis (TRIPOD): explanation and elaboration. Ann Intern Med. 2015;162(1):W1–73.

17. Hunt LP, Ben-Shlomo Y, Clark EM, Dieppe P, Judge A, MacGregor AJ, et al. 45-day mortality after 467,779 knee replacements for osteoarthritis from the National Joint Registry for England and Wales: an observational study. Lancet. 2014;384(9952):1429–36.

